# Pathways linking pulse pressure to dementia in adults with Down syndrome

**DOI:** 10.1101/2023.10.26.23297625

**Authors:** Batool Rizvi, Patrick J. Lao, Mithra Sathishkumar, Lisa Taylor, Nazek Queder, Liv McMillan, Natalie Edwards, David B. Keator, Eric Doran, Christy Hom, Dana Nguyen, H. Diana Rosas, Florence Lai, Nicole Schupf, Jose Gutierrez, Wayne Silverman, Ira T. Lott, Mark Mapstone, Donna M. Wilcock, Elizabeth Head, Michael A. Yassa, Adam M. Brickman

## Abstract

Individuals with Down syndrome (DS) are less likely to have hypertension than neurotypical adults. However, whether blood pressure measures are associated with brain health and clinical outcomes in this population has not been studied in detail. Here, we assessed whether pulse pressure is associated with markers of cerebrovascular disease, entorhinal cortical atrophy, and diagnosis of dementia in adults with DS. Participants with DS from the Biomarkers of Alzheimer’s Disease in Adults with Down Syndrome study (ADDS; n=195, age=50.6±7.2 years, 44% women, 18% diagnosed with dementia) were included. Higher pulse pressure was associated with greater global, parietal, and occipital WMH volume. Pulse pressure was not related to enlarged PVS, microbleeds, infarcts, entorhinal cortical thickness, or dementia diagnosis. However, in a serial mediation model, we found that pulse pressure was indirectly related to dementia diagnosis through parieto-occipital WMH and, subsequently through entorhinal cortical thickness. Higher pulse pressure may be a risk factor for dementia in people with DS by promoting cerebrovascular disease, which in turn affects neurodegeneration. Pulse pressure is an important determinant of brain health and clinical outcomes in individuals with Down syndrome despite the low likelihood of frank hypertension.

## INTRODUCTION

As individuals with Down syndrome (DS) reach 50 years of age, nearly all have sufficient beta-amyloid and tau pathology to meet the pathological criteria for Alzheimer’s disease (AD) ^1-5^. Despite the inevitable accumulation of beta-amyloid and tau pathology in adults with DS, there is some variability in the age of onset of clinical symptoms of dementia and in the severity and the course of decline of clinical symptoms ^2,4,6^. Moreover, a small subset of adults with DS do not develop dementia symptoms in their lifetime ^7^. The factors that account for the variability in clinical AD onset and course among adults with DS are poorly understood. Although AD pathogenesis in the context of DS is typically attributed to a single pathway model (i.e., the amyloid cascade hypothesis) ^8-10^, and the clinical course is typically framed according to the amyloid-tau-neurodegeneration (ATN) biomarker classification scheme ^11,12^, there is increasing evidence of the contribution of cerebrovascular disease to both clinical and pathogenic progression ^13-16^. Despite relatively low prevalence of classical vascular risk factors ^17,18^, individuals with DS have cerebrovascular abnormalities on magnetic resonance imaging (MRI), including white matter hyperintensities (WMH), enlarged perivascular spaces (PVS), microbleeds, and infarcts, which increase across AD-related clinical diagnoses ^19^.

In the neurotypical population the presence and severity of vascular risk factors, particularly high blood pressure (hypertension), lead to cerebrovascular disease, which in turn, is associated with neurodegeneration, cognitive decline, and risk and progression of clinical AD ^20-22^. These relationships have not been examined systematically among adults with DS, likely due to the low prevalence of vascular risk factors in people with DS ^18^. Individuals with DS have lower blood pressure than individuals from the neurotypical population ^23-25^, as well as lower prevalence and incidence of hypertension ^26^. The physiological mechanisms of these observations are not well understood. Nonetheless, it is possible that increases in blood pressure measurements may be associated with poorer brain outcomes among older adults with DS even if those measurements are within the normal range for the neurotypical population. In support of this hypothesis, studies in the neurotypical population found that elevated blood pressure even in non-hypertensive adults is associated with worse cognition ^27,28^.

In the current study, we examined the association of common clinical blood pressure-related measurements of vascular health with MRI markers of small vessel disease or dysfunction, entorhinal cortical thickness, and clinical dementia status in older adults with DS. Systolic blood pressure (SBP) measures the force of the heart exerting on artery walls at each beat, while diastolic blood pressure (DBP) measures the force of the heart exerting on the artery walls in between beats, and is a measure of peripheral resistance. Pulse pressure (PP), or the difference between systolic and diastolic blood pressure, is an indicator of arterial stiffness. Mean arterial pressure (MAP) is the average arterial pressure during one cardiac cycle. ^29^. In the neurotypical population, hypertension is related to increased risk for stroke, WMH, infarcts, and AD related pathology ^30^. Increased pulse pressure can lead to atherosclerosis even in normotensive individuals ^31^, which is associated with WMH ^32,33^. Higher pulse pressure is also associated with medial temporal lobe atrophy in AD ^34^. Compared with other measures of blood pressure, pulse pressure is more tightly linked to AD pathology ^35,36^. Higher pulse pressure can affect cerebral blood flow, gray matter and white matter integrity, increasing the risk for dementia ^37,38^.

Given the documented stronger associations of clinical outcomes with pulse pressure than other blood pressure indicators ^35,39^, we focused primarily on this measure in the current analyses. We assessed the associations of these blood pressure measures with MRI markers of cerebrovascular disease, entorhinal cortical thickness, and clinical diagnosis of dementia. Previously, we showed that parietal and occipital lobe WMH are associated with medial temporal lobe atrophy in community-dwelling older adults ^40-42^. Prior work also demonstrated that greater parietal and occipital lobe WMH burden is predictive of a diagnosis of dementia in adults with DS and in autosomal dominant AD ^19,43^. Based on these observations, we hypothesized that increased pulse pressure is related to greater parietal and occipital WMH, which in turn is related to lower entorhinal cortical thickness, and subsequently to dementia.

## MATERIALS AND METHODS

### Participants

This study included participants from the Biomarkers of Alzheimer’s Disease in Adults with Down Syndrome (ADDS; U01 AG051412) study, which characterizes the development of AD dementia among older adults with DS. Participants were enrolled at multiple sites, including Columbia University/New York State Institute for Basic Research in Developmental Disabilities (n=54), Massachusetts General Hospital/Harvard Medical School (n=75), and University of California, Irvine (n=66). Participants whose blood pressure was measured during their baseline visit were included in the analyses (N=195).

The study was approved by the institutional review boards of the participating institutions, and written informed consent was obtained from the participants and/or their legal guardian or legally authorized representative. We received assent from every participant before every procedure.

### Blood Pressure Measures

Blood pressure, including systolic and diastolic blood pressure was assessed within three months of the MRI visit. Blood pressure was assessed within a single measurement while the participant was seated. Pulse pressure was derived by taking the difference between systolic blood pressure and diastolic blood pressure: PP = SBP - DBP. Mean arterial pressure was calculated using the following equation: MAP = (SBP + 2(DBP)) / 3

### Clinical Diagnosis

As part of the diagnostic procedure, participants underwent neuropsychological testing to assess cognition in domains typically affected by AD. Study personnel reviewed clinical charts, conducted interviews with knowledgeable informants, and conducted a standardized clinical and neurological examination. A consensus panel of clinicians, expert in the diagnosis of dementia in individuals with DS, assigned a final diagnosis based on the information collected, which did not include any biomarker data ^44^. One of four AD-related diagnoses was assigned to each participant: cognitively stable (CS), mild cognitive impairment-DS (MCI-DS), possible AD dementia, and definite AD-dementia. For our analyses, we categorized individuals into two groups: with AD dementia (including possible and definite AD dementia) or without dementia (including CS or MCI-DS).

### MRI imaging

MRI scans (n=145) were acquired on a Siemens Prisma 3T at Columbia University (n=30) and MGH (n=58) or a Philips Achieva 3T at UC Irvine (n=57). The Alzheimer’s Disease Neuroimaging Initiative MRI protocol was used at all sites (T1-weighted scan: repetition time (TR)/echo time (TE)/inversion time (TI): 2300/2.96/900ms; voxel size: 1x1x1mm^3^; T2-weighted fluid attenuated inversion recovery (FLAIR) scan (TR/TE/TI: 5000/386/1800ms; voxel size: 0.4x0.4x0.9mm^3^); and a T2-*-weighted gradient echo (GRE) scan (TR/TE: 650/20ms; voxel size: 0.8x0.8x4mm^3^) or susceptibility weighted image (SWI: TRE/TE: 27/30ms; voxel size: 0.9x0.9x1.5 mm^3^)). Total and regional (by cerebral lobe) WMH volumes were quantified from T2-weighted FLAIR scans, using a previously described method ^19,45^. Parietal and occipital WMH were summed to derive parieto-occipital WMH volume. Enlarged perivascular spaces (PVS) were visually rated as hypointensities on T1 scans across 13 brain regions and rated from 0 to 2 based on FLAIR characteristics. These ratings were then summed for a global score ranging from 0 (no enlarged PVS in any region) to 26 (severe enlarged PVS in all regions) ^46^. Infarcts were visually counted on FLAIR scans as discrete hypointense lesions (> 5 mm) with partial or complete hyperintense ring and confirmed on T1 scans (hypointense areas) and were globally scored as present or not. Microbleeds were counted by visual inspection, as hypointense round or ovoid lesions on GRE or SWI, and were globally scored as present or not.

### Entorhinal cortical thickness

Each participant’s T1-weighted image was processed with FreeSurfer v.6.0 (*http://surfer.nmr.mgh.harvard.edu/)* to derive left and right entorhinal cortical thickness. Left and right entorhinal cortical thickness were averaged when included in the mediation models.

### Statistical Analysis

General linear models were used to test the association of pulse pressure with regional WMH, enlarged PVS score, and entorhinal cortical thickness. We used logistic regression models to test the association between pulse pressure and the presence of microbleeds, infarcts, and dementia. In a mediation model, we first tested the hypothesis that high pulse pressure is associated with dementia through parieto-occipital WMH. Additionally, in a serial mediation model, we tested the mechanistic hypothesis that high pulse pressure is related to increased dementia through parieto-occipital WMH and lower entorhinal cortical thickness. We used the SPSS PROCESS macro v.3.5 (processmacro.org), and applied a 95% confidence interval (CI) with 5000 bootstrap samples for both mediation analyses. All analyses were adjusted for age and sex. In a sensitivity analysis, we tested whether associations remained after removing participants who were treated for either hypertension or hypotension. Associations with SBP, DBP and MAP were reported in supplementary results.

## RESULTS

One-hundred ninety-five participants were included. Table 1 displays summary characteristics, including demographic, clinical, and imaging variables, and the sample size associated with each variable. About 17% of the participants with DS were characterized as having high blood pressure (hypertension; SBP ≥130 and/or DBP ≥ 80 mmHg), while around 28% demonstrated low blood pressure (hypotension; SBP < 90 and/or DBP < 60 mmHg).

**Table 1.**
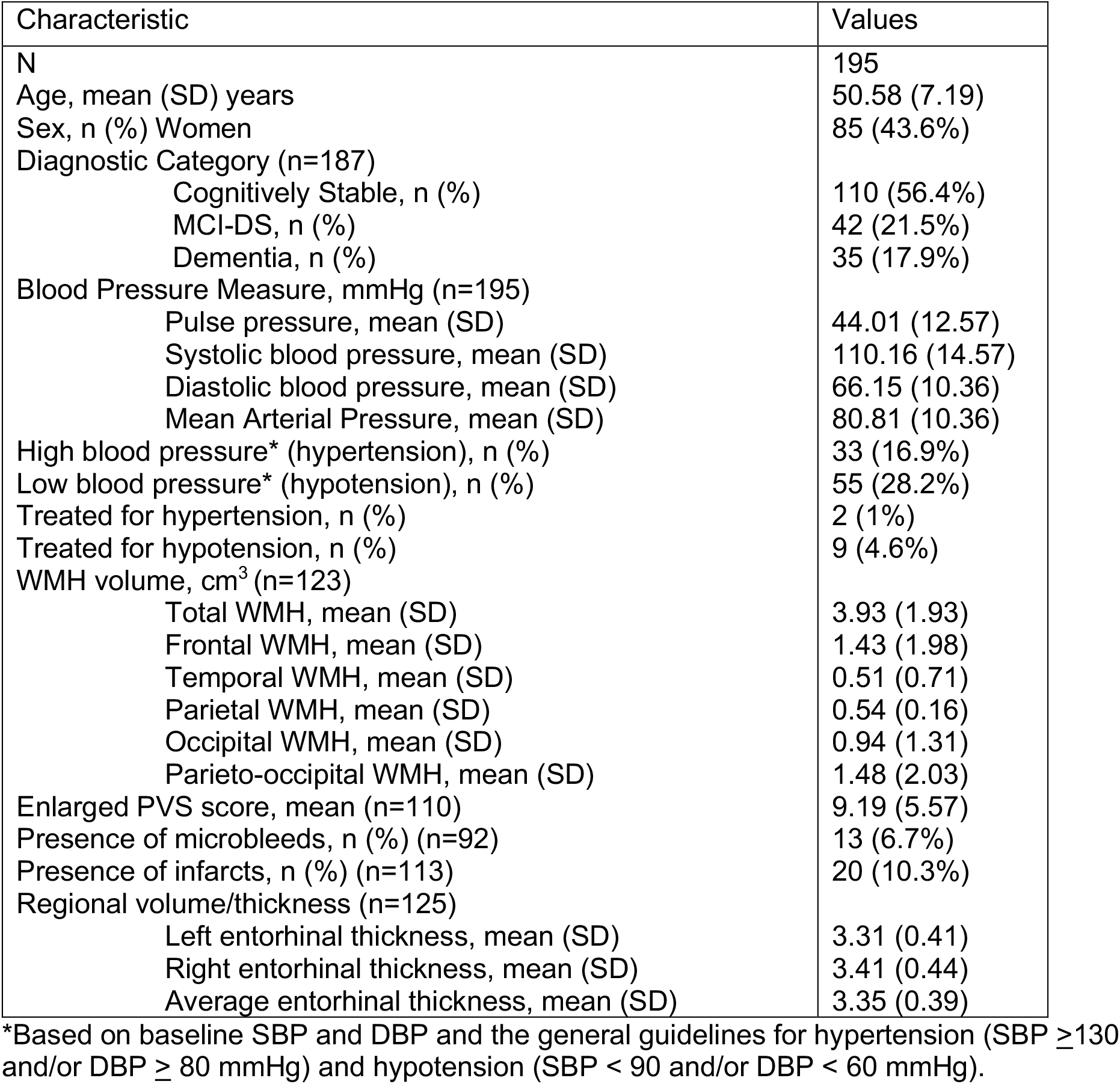

### 1. Association of pulse pressure with cerebrovascular imaging markers

Higher pulse pressure was associated with greater global WMH, parietal WMH, and occipital WMH (see Table 2 and Figure 2A, 2B). In a sensitivity analysis, after removing participants who were treated for either hypotension or hypertension, the findings were similar with the exception that higher pulse pressure was additionally associated with greater temporal WMH (see Supplementary Table 1). Higher systolic blood pressure was associated occipital WMH and marginally with temporal WMH (see Supplementary Table 2). Diastolic blood pressure and MAP were not associated with global or regional WMH volume (see Supplementary Table 2).

**Table 2.**

| BP Measure | Outcome Measure | *B* | $\beta$ | p-value | 95% CI |
| --- | --- | --- | --- | --- | --- |
| Pulse pressure | Global WMH | 0.071 | 0.191 | 0.032* | (0.006, 0.135) |
|  | Frontal WMH | 0.016 | 0.100 | 0.264 | (-0.012, 0.045) |
|  | Temporal WMH | 0.010 | 0.174 | 0.059 | (0.000, 0.021) |
|  | Parietal WMH | 0.015 | 0.204 | 0.023* | (0.002, 0.027) |
|  | Occipital WMH | 0.022 | 0.208 | 0.017* | (0.004, 0.041) |
|  | Cortical microbleeds<sup>λ</sup> | -0.009 | 0.991 | 0.704 | (0.946, 1.038) |
|  | Perivascular spaces | 0.075 | 0.168 | 0.070 | (-0.006, 0.156) |
|  | Infarcts<sup>λ</sup> | 0.020 | 1.020 | 0.327 | (0.980, 1.061) |
|  | Left EC thickness | 0.003 | 0.102 | 0.251 | (-0.002, 0.009) |
|  | Right EC thickness | 0.001 | 0.036 | 0.695 | (-0.005, 0.007) |
|  | Clinical Dementia Diagnosis | 0.015 | 1.015 | 0.322 | (0.986, 1.045) |

Primary regression model: Outcome measure ~ BP measure + age + intercept.
\* Significant with a p-value < 0.05
<sup>λ</sup> For these dichotomous dependent variables, logistic regression was applied. $\beta$ = Exponential ( $\beta$ ) and 95%CI = 95% CI for Exponential ( $\beta$ )

**Figure 1.**
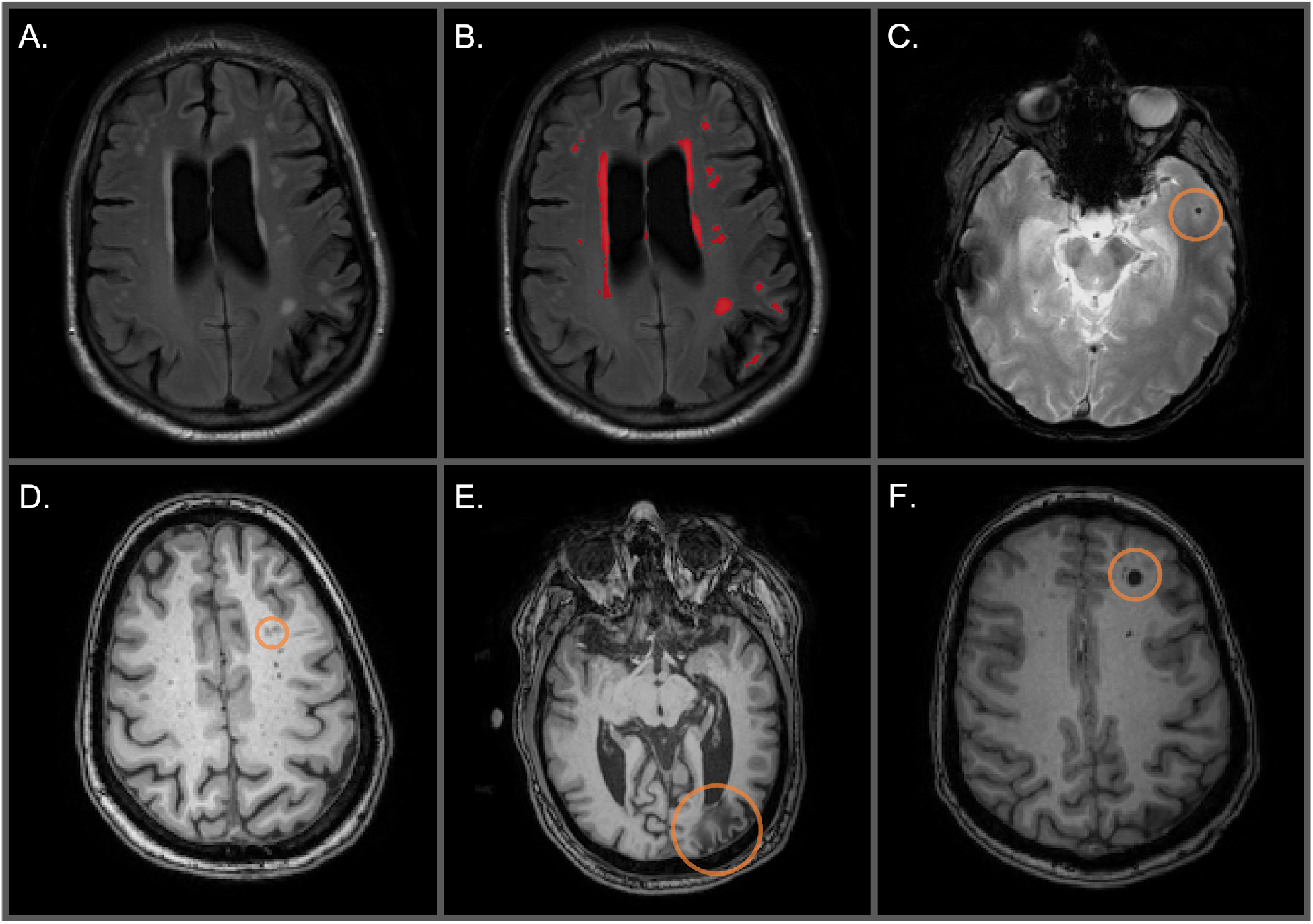
**A**. Unlabeled WMH on an axial FLAIR image. **B**. Labeled WMH in red on an axial FLAIR image **C**. Cortical microbleed circled on an axial T2*-GRE. **D**. Perivascular spaces circled on an axial T1-weighed image. **E**. Distal cortical infarct circled on an axial T1-weighted image. **F**. Deep infarct circled on an axial T1-weighted image.

**Figure 2.**
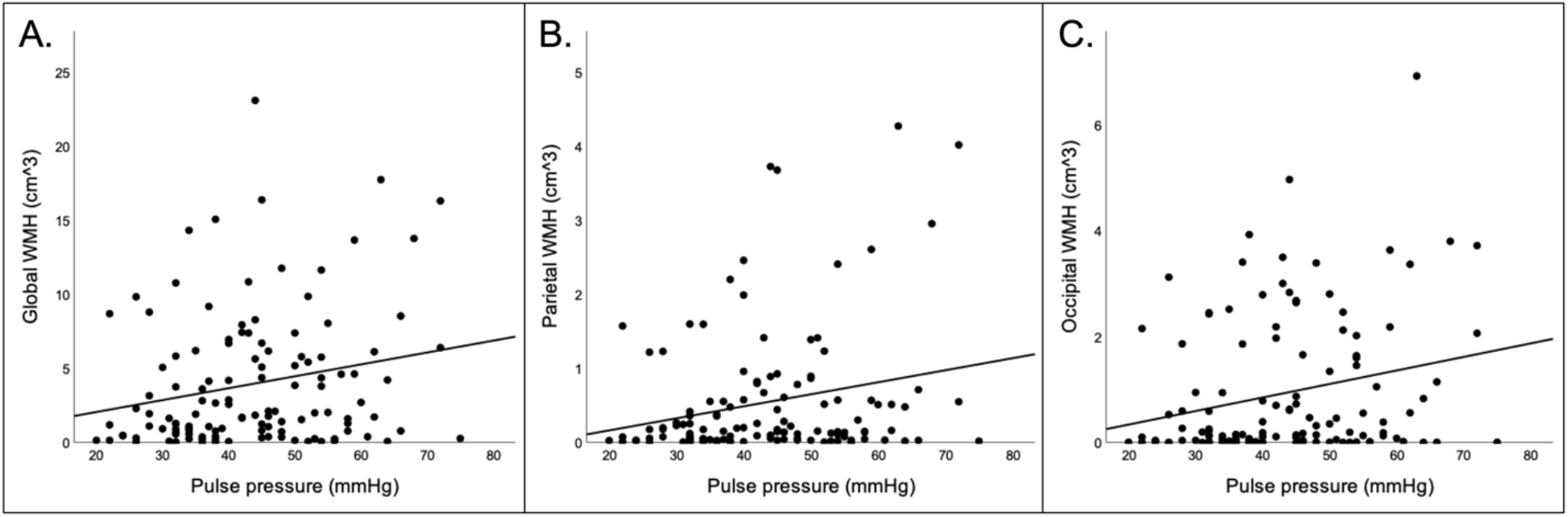
Scatterplots displaying associations between: A) pulse pressure and global WMH; B) pulse pressure and parietal WMH; C) pulse pressure and occipital WMH.

Pulse pressure and systolic blood pressure were not associated with enlarged perivascular spaces, microbleeds, or infarcts in primary (see Table 2) or sensitivity analyses (see Supplementary Table 1). Diastolic blood pressure and MAP were also not associated with enlarged perivascular spaces, microbleeds, or infarcts (See Supplementary Table 2).

### 2. Direct and indirect effects of pulse pressure on dementia

Higher pulse pressure was not associated with left or right entorhinal cortical thickness (see Table 2). Similarly, systolic blood pressure, diastolic blood pressure, and mean arterial pressure were not related to left or right entorhinal cortical thickness (see Supplementary Table 2). Higher pulse pressure was also not associated with clinical dementia status (see Table 2). Similarly, systolic blood pressure, diastolic pressure, and mean arterial pressure were not related to dementia diagnosis. The lack of direct associations prompted us to consider indirect pathways in which pulse pressure would affect neurodegeneration and clinical status through cerebrovascular pathology.

In a simple mediation analysis, we found that there was an indirect (mediating) effect of higher pulse pressure on diagnosis of dementia through greater parieto-occipital WMH (see Figure 3). We next tested in a serial mediation whether pulse pressure would lead to greater parieto-occipital WMH, thereby lowering entorhinal cortical thickness, and subsequently lead to increased dementia (see Figure 4). There was an indirect effect by which greater parieto-occipital WMH and lower entorhinal cortical thickness serially mediated the association of higher pulse pressure on increased dementia (see Figure 4).

**Figure 3.**
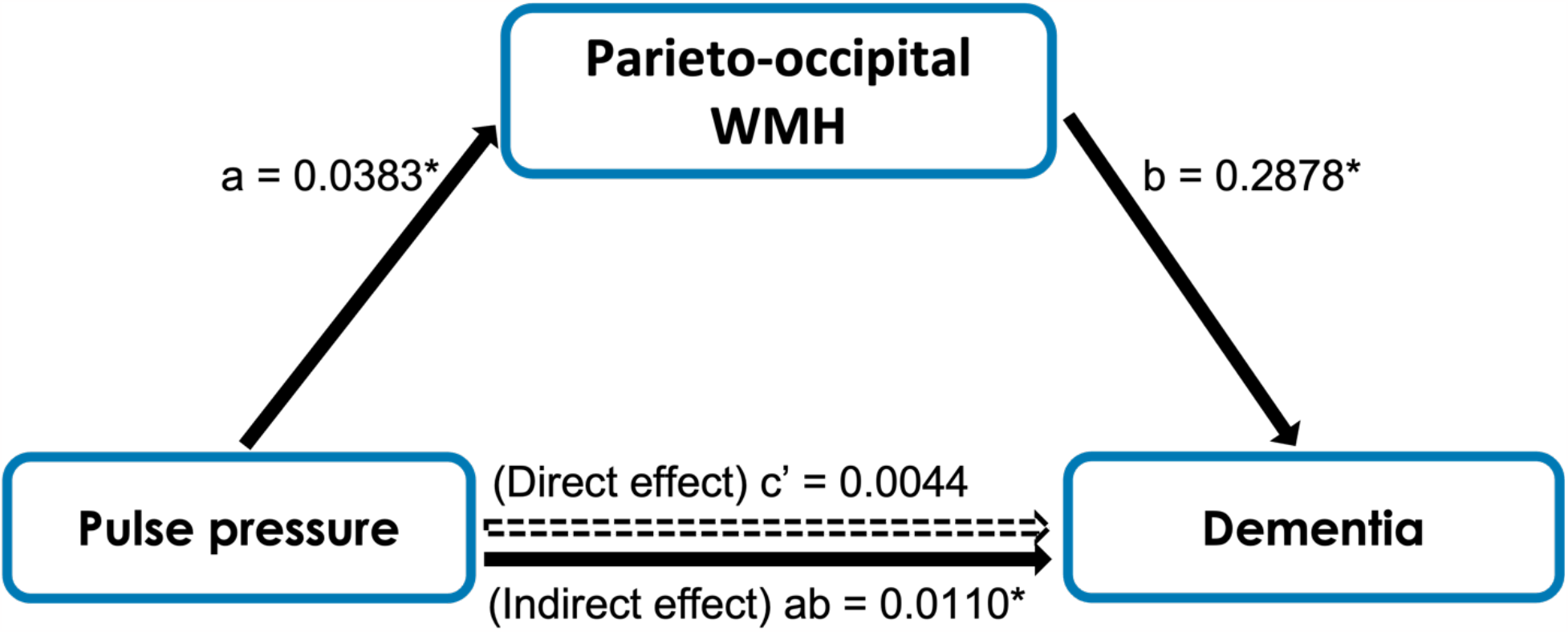
Mediation model of pulse pressure, parieto-occipital WMH, and status of dementia. An indirect effect was found where parieto-occipital WMH mediated the effect of pulse pressure on the frequency of dementia. * Indicates statistical significance. Path a: *b* = 0.0383, p = 0.0611 Path b: *b* = 0.2868, p = 0.0221 Path c’ (Direct effect): *b* = 0.0044, 95% CI (-0.0439, 0.0527) Path ab (Indirect effect): *b* = 0.0110, 95% CI (0.0004, 0.0297)

**Figure 4.**
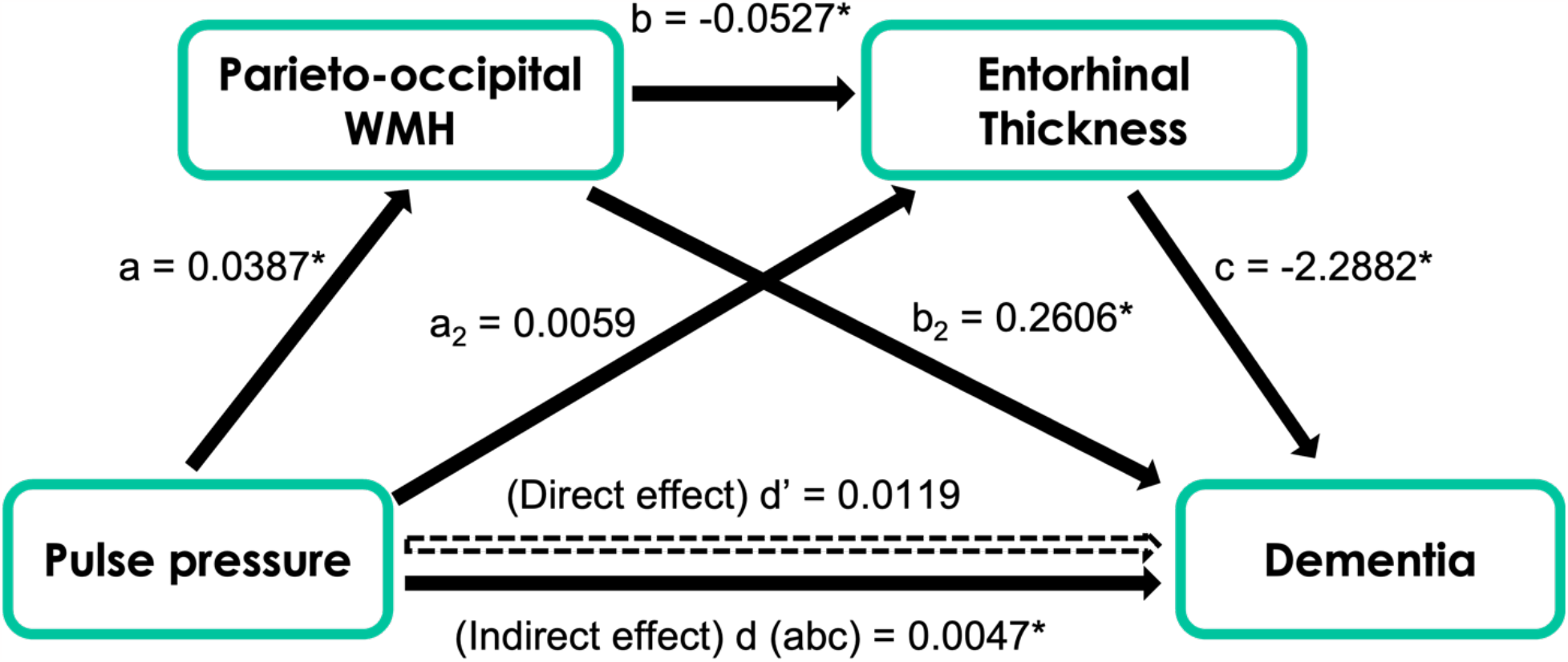
Mediation model of pulse pressure, parieto-occipital WMH, entorhinal cortical thickness, and dementia. An indirect effect was found where parieto-occipital WMH and entorhinal cortical thickness serially mediated the effect of pulse pressure on frequency of dementia. * Indicates statistical significance. Path a: *b* = 0.0387, p = 0.0134 Path b: *b* = -0.0527, p = 0.0055 Path c: *b* = -2.2882, p = 0.0173 Path d’ (Direct effect): *b* = 0.0119, 95% CI (-0.0437, 0.0675) Path d (abc; indirect effect): *b* = 0.0047, 95% CI (0.0003, 0.0179)

## DISCUSSION

Higher pulse pressure was associated with greater global, parietal, and occipital WMH in older adults with DS, but not with other markers of cerebrovascular disease. Pulse pressure was not directly associated with entorhinal cortical thickness or dementia status. However, higher pulse pressure was indirectly associated with diagnosis of dementia, through parieto-occipital WMH in a simple mediation. Furthermore, in a serial mediation, which allowed us to formally test the mechanistic framework we hypothesized linking pulse pressure to dementia, the relationship between pulse pressure and dementia status was sequentially mediated by both parieto-occipital WMH and lower entorhinal cortical thickness, suggesting that increased pulse pressure leads to cerebrovascular disease, which promotes regional atrophy and subsequent dementia.

Blood pressure has not been studied comprehensively in relation to brain or any clinical marker associated with AD in adults with DS. To our knowledge, only one previous study examined blood pressure in DS, and found that blood pressure did not increase with age in people with DS despite observed age-associated increases in neurotypical participants^23^. The current relationship of pulse pressure with cerebrovascular disease was specific to only white matter pathology, which was not unexpected. We did not find any association between pulse pressure or systolic blood pressure with enlarged perivascular spaces, microbleeds, or infarcts, suggesting that that WMH, a marker of chronic and subtle small vessel disease ^47^, is more sensitive to the impact of relatively higher pulse pressure. The other cerebrovascular imaging markers may indicate other mechanistic processes, including impaired glymphatic clearance resulting in enlarged perivascular spaces ^48^, cerebral amyloid angiopathy that is more frequent in the brains of people with DS and associated with microbleeds ^13,49,50^, and large artery disease leading to infarcts ^51^.

The lack of direct association of pulse pressure with dementia may be explained by the mechanistic chain of events occurring prior to the onset of dementia. For example, higher pulse pressure may directly induce more cerebrovascular changes as observed in our study rather than act on neurodegenerative processes directly, which may lead to a cascade of events, such as lower entorhinal thickness and subsequent clinical impairment. This possibility is highlighted in the serial mediation model, in which parieto-occipital WMH and entorhinal cortical thickness mediated the association between higher pulse pressure and increased dementia. The association between parieto-occipital WMH and lower entorhinal thickness is supported by our previous work ^40 41^and further investigation is needed on whether this association is due to the lowered perfusion of the posterior cerebral artery supplying both posterior and medial temporal lobe regions.

This study has some limitations. Due to the cross-sectional nature of the current study, we could not investigate the temporality or causality of the observed relationships. Based on a number of studies highlighting the role of blood pressure as a risk factor for age- and AD-related changes in the neurotypical population (see reviews ^30,52,53^), it is more likely that higher blood pressure and pulse pressure precedes the pathophysiological markers. Secondly, the adults with DS included in the study were predominantly non-Hispanic White. In the neurotypical aging population, higher blood pressure is more prevalent among racially and ethnically minoritized populations ^54^, leading to further health disparities. Another future direction is to examine markers other than pulse pressure that can more accurately capture arterial stiffness in adults with DS, such as with intracranial pulse wave velocity ^55^. Another area of future work would be to examine how blood pressure measures are associated with AD related pathology, such as by amyloid and tau PET imaging.

To summarize, this work demonstrates that higher pulse pressure is associated with global and more posterior WMH, including parietal and occipital WMH and indirectly associated with dementia through greater parieto-occipital WMH burden and lower entorhinal cortical thickness. This work highlights the impact of elevated pulse pressure and that the range of blood pressure values provide novel information within individuals with DS. Monitoring and targeting blood pressure through nonpharmacological or pharmacological approaches may be helpful for people with DS.

## Data Availability

Data in the current study came from the Alzheimer's Biomarker Consortium - Down Syndrome (ABC-DS) study. Data from the ABC-DS can be requested here:
https://www.nia.nih.gov/research/abc-ds#data

## SUPPLEMENT

**Supplementary Table 1.**
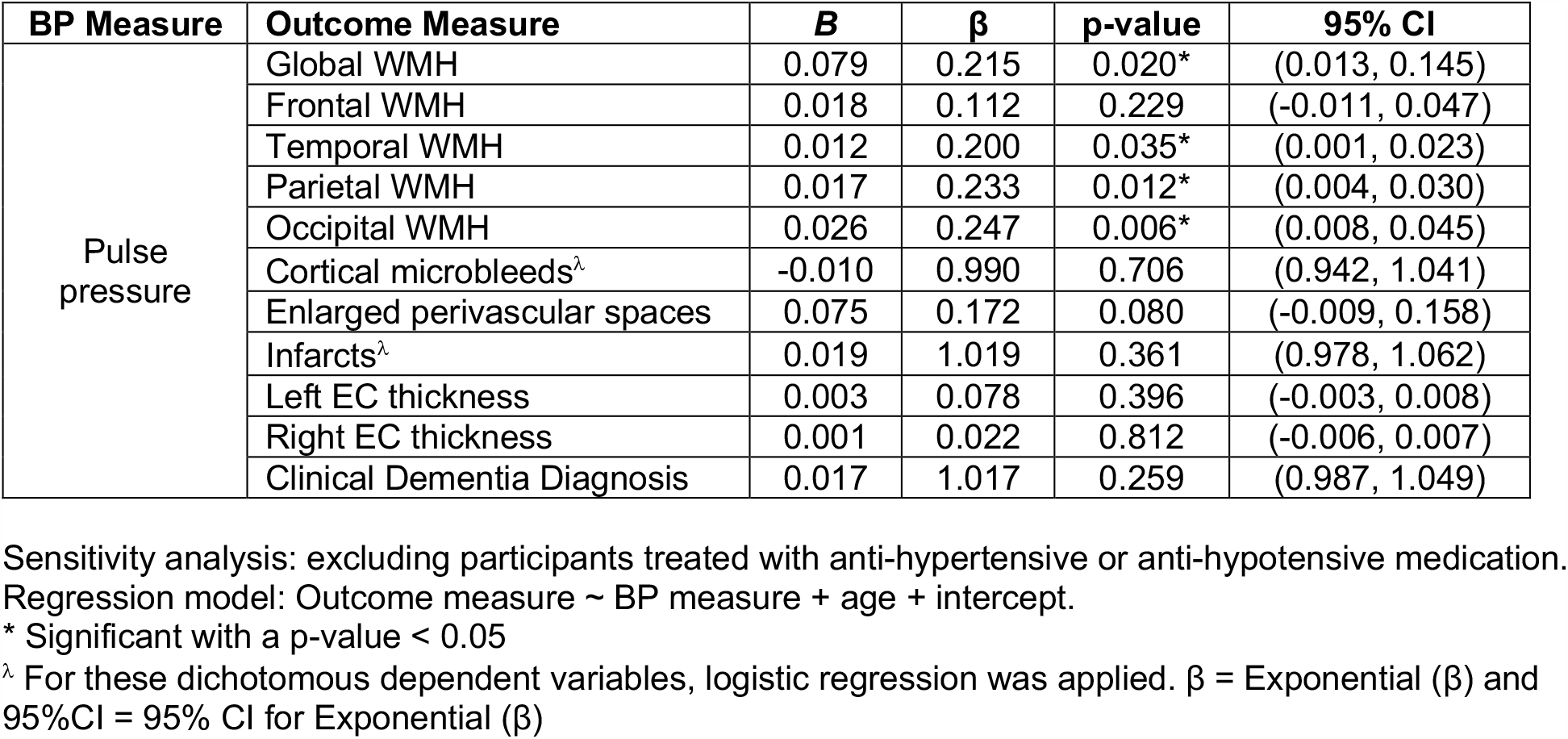

**Supplementary Table 2.**
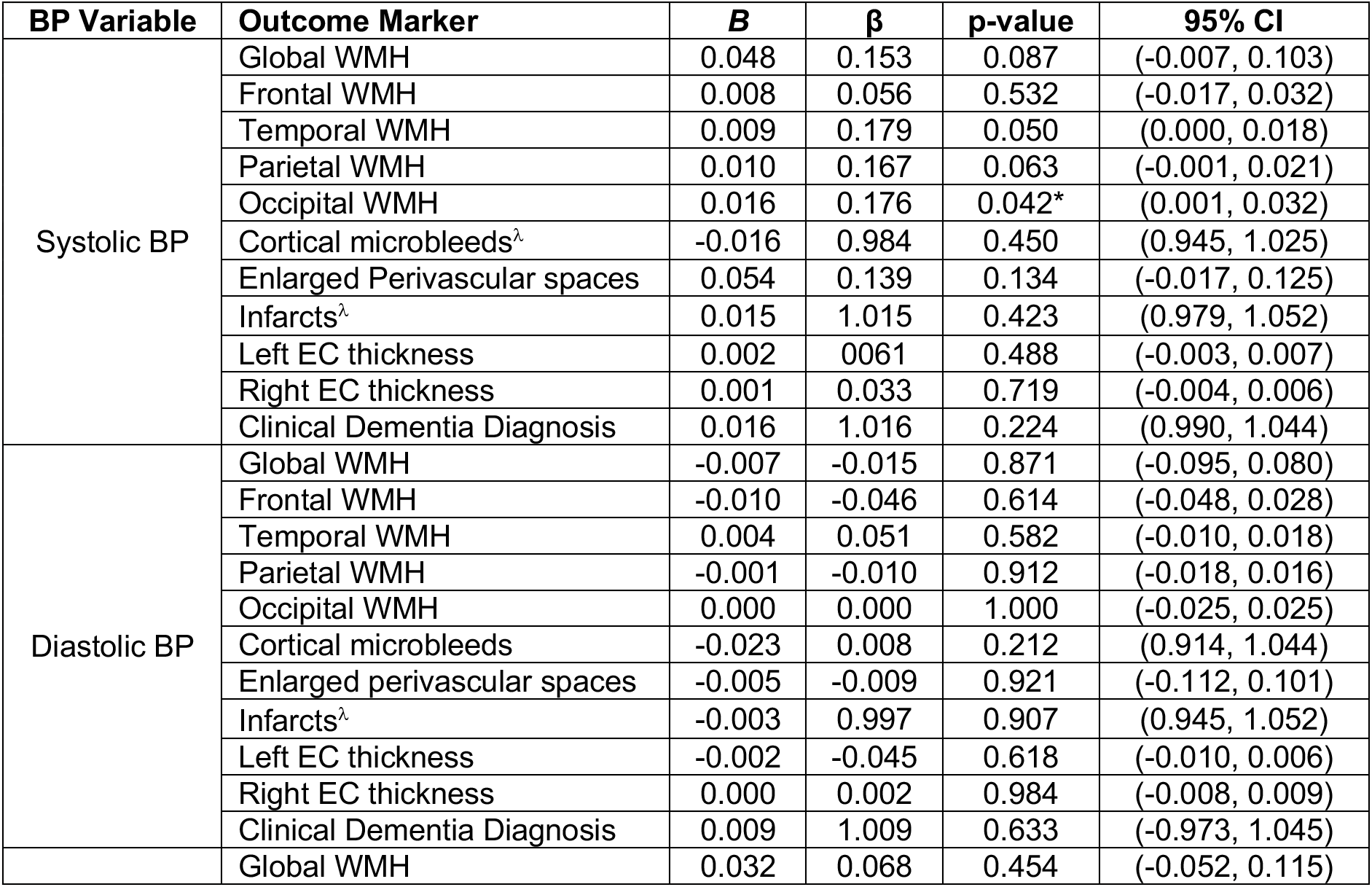
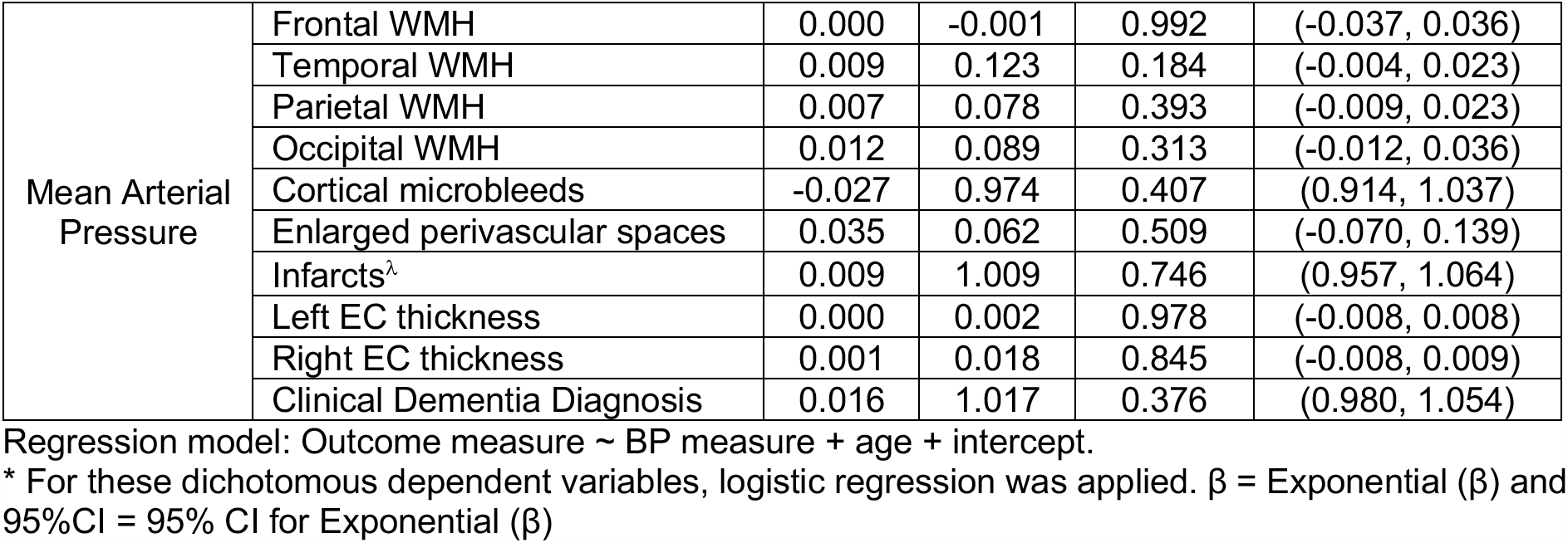

### Funding

This research was supported by grants from the National Institutes of Health (NIA U01 AG051412, U19 AG068054, RF1 AG079519).

## References

1. Head E, Lott IT, Wilcock DM, Lemere CA. Aging in Down Syndrome and the Development of Alzheimer’s Disease Neuropathology. Curr Alzheimer Res. 2016;13(1):18–29.

2. Lott IT, Head E. Dementia in Down syndrome: unique insights for Alzheimer disease research. Nature reviews Neurology. 2019;15(3):135–147.

3. Fortea J, Vilaplana E, Carmona-Iragui M, et al. Clinical and biomarker changes of Alzheimer’s disease in adults with Down syndrome: a cross-sectional study. Lancet. 2020;395(10242):1988–1997.

4. Fortea J, Zaman SH, Hartley S, Rafii MS, Head E, Carmona-Iragui M. Alzheimer’s disease associated with Down syndrome: a genetic form of dementia. Lancet Neurol. 2021;20(11):930–942.

5. McCarron M, McCallion P, Reilly E, Dunne P, Carroll R, Mulryan N. A prospective 20-year longitudinal follow-up of dementia in persons with Down syndrome. J Intellect Disabil Res. 2017;61(9):843–852.

6. Sinai A, Mokrysz C, Bernal J, et al. Predictors of Age of Diagnosis and Survival of Alzheimer’s Disease in Down Syndrome. Journal of Alzheimer’s disease : JAD. 2018;61(2):717–728.

7. Head E, Powell D, Gold BT, Schmitt FA. Alzheimer’s Disease in Down Syndrome. Eur J Neurodegener Dis. 2012;1(3):353–364.

8. Glenner GG, Wong CW. Alzheimer’s disease and Down’s syndrome: sharing of a unique cerebrovascular amyloid fibril protein. Biochem Biophys Res Commun. 1984;122(3):1131–1135.

9. Hardy JA, Higgins GA. Alzheimer’s disease: the amyloid cascade hypothesis. Science (New York, NY). 1992;256(5054):184–185.

10. Head E, Helman AM, Powell D, Schmitt FA. Down syndrome, beta-amyloid and neuroimaging. Free Radic Biol Med. 2018;114:102–109.

11. Rafii MS, Ances BM, Schupf N, et al. The AT(N) framework for Alzheimer’s disease in adults with Down syndrome. Alzheimers Dement (Amst). 2020;12(1):e12062.

12. Jack CR, Jr., Bennett DA, Blennow K, et al. NIA-AA Research Framework: Toward a biological definition of Alzheimer’s disease. Alzheimer’s & dementia : the journal of the Alzheimer’s Association. 2018;14(4):535–562.

13. Head E, Phelan MJ, Doran E, et al. Cerebrovascular pathology in Down syndrome and Alzheimer disease. Acta neuropathologica communications. 2017;5(1):93.

14. Kalaria RN. Cerebrovascular disease and mechanisms of cognitive impairment: evidence from clinicopathological studies in humans. Stroke. 2012;43(9):2526–2534.

15. Vemuri P, Knopman DS. The role of cerebrovascular disease when there is concomitant Alzheimer disease. Biochim Biophys Acta. 2016;1862(5):952–956.

16. Laing KK, Simoes S, Baena-Caldas GP, et al. Cerebrovascular disease promotes tau pathology in Alzheimer’s disease. Brain Commun. 2020;2(2):fcaa132.

17. Rizvi B, Head E, Brickman AM. The role of cerebrovascular disease in aging and Alzheimer’s disease among people with Down syndrome. The Neurobiology of Aging and Alzheimer Disease in Down Syndrome. 2022:63–73.

18. Wilcock DM, Schmitt FA, Head E. Cerebrovascular contributions to aging and Alzheimer’s disease in Down syndrome. Biochim Biophys Acta. 2016;1862(5):909–914.

19. Lao PJ, Gutierrez J, Keator D, et al. Alzheimer-Related Cerebrovascular Disease in Down Syndrome. Annals of neurology. 2020;88(6):1165–1177.

20. Qiu C, Winblad B, Fratiglioni L. The age-dependent relation of blood pressure to cognitive function and dementia. Lancet Neurol. 2005;4(8):487–499.

21. Sierra C, Lopez-Soto A, Coca A. Connecting cerebral white matter lesions and hypertensive target organ damage. J Aging Res. 2011;2011:438978.

22. Sierra C. Hypertension and the Risk of Dementia. Front Cardiovasc Med. 2020;7:5.

23. Morrison RA, McGrath A, Davidson G, Brown JJ, Murray GD, Lever AF. Low blood pressure in Down’s syndrome, A link with Alzheimer’s disease? Hypertension. 1996;28(4):569–575.

24. Richards BW, Enver F. Blood pressure in Down’s syndrome. J Ment Defic Res. 1979;23(2):123–135.

25. Rodrigues AN, Coelho LC, Goncalves WL, et al. Stiffness of the large arteries in individuals with and without Down syndrome. Vasc Health Risk Manag. 2011;7:375–381.

26. Alexander M, Petri H, Ding Y, Wandel C, Khwaja O, Foskett N. Morbidity and medication in a large population of individuals with Down syndrome compared to the general population. Dev Med Child Neurol. 2016;58(3):246–254.

27. Knecht S, Wersching H, Lohmann H, et al. High-normal blood pressure is associated with poor cognitive performance. Hypertension. 2008;51(3):663–668.

28. Jennings JR, Muldoon MF, Ryan C, et al. Prehypertensive Blood Pressures and Regional Cerebral Blood Flow Independently Relate to Cognitive Performance in Midlife. J Am Heart Assoc. 2017;6(3).

29. DeMers D, Wachs D. Physiology, Mean Arterial Pressure. In: StatPearls. Treasure Island (FL) ineligible companies. Disclosure: Daliah Wachs declares no relevant financial relationships with ineligible companies. 2023.

30. Skoog I, Gustafson D. Update on hypertension and Alzheimer’s disease. Neurological research. 2006;28(6):605–611.

31. Zakopoulos NA, Lekakis JP, Papamichael CM, et al. Pulse pressure in normotensives: a marker of cardiovascular disease. Am J Hypertens. 2001;14(3):195–199.

32. Safar ME. Pulse pressure in essential hypertension: clinical and therapeutical implications. J Hypertens. 1989;7(10):769–776.

33. Zang J, Shi J, Liang J, et al. Pulse Pressure, Cognition, and White Matter Lesions: A Mediation Analysis. Front Cardiovasc Med. 2021;8:654522.

34. Korf ES, Scheltens P, Barkhof F, de Leeuw FE. Blood pressure, white matter lesions and medial temporal lobe atrophy: closing the gap between vascular pathology and Alzheimer’s disease? Dementia and geriatric cognitive disorders. 2005;20(6):331–337.

35. Nation DA, Delano-Wood L, Bangen KJ, et al. Antemortem pulse pressure elevation predicts cerebrovascular disease in autopsy-confirmed Alzheimer’s disease. Journal of Alzheimer’s disease : JAD. 2012;30(3):595–603.

36. Weigand AJ, Macomber AJ, Walker KS, et al. Interactive Effects of Pulse Pressure and Tau Imaging on Longitudinal Cognition. Journal of Alzheimer’s disease : JAD. 2022;89(2):633–640.

37. Badji A, Sabra D, Bherer L, Cohen-Adad J, Girouard H, Gauthier CJ. Arterial stiffness and brain integrity: A review of MRI findings. Ageing Res Rev. 2019;53:100907.

38. Waldstein SR, Rice SC, Thayer JF, Najjar SS, Scuteri A, Zonderman AB. Pulse pressure and pulse wave velocity are related to cognitive decline in the Baltimore Longitudinal Study of Aging. Hypertension. 2008;51(1):99–104.

39. Safar ME, Nilsson PM, Blacher J, Mimran A. Pulse pressure, arterial stiffness, and end-organ damage. Curr Hypertens Rep. 2012;14(4):339–344.

40. Rizvi B, Narkhede A, Last BS, et al. The effect of white matter hyperintensities on cognition is mediated by cortical atrophy. Neurobiol Aging. 2018;64:25–32.

41. Rizvi B, Lao PJ, Chesebro AG, et al. Association of Regional White Matter Hyperintensities With Longitudinal Alzheimer-Like Pattern of Neurodegeneration in Older Adults. JAMA Netw Open. 2021;4(10):e2125166.

42. Rizvi B, Sathishkumar M, Kim S, et al. Posterior white matter hyperintensities are associated with reduced medial temporal lobe subregional integrity and long-term memory in older adults. Neuroimage Clin. 2023;37:103308.

43. Lee S, Viqar F, Zimmerman ME, et al. White matter hyperintensities are a core feature of Alzheimer’s disease: Evidence from the dominantly inherited Alzheimer network. Annals of neurology. 2016;79(6):929–939.

44. Krinsky-McHale SJ, Zigman WB, Lee JH, et al. Promising outcome measures of early Alzheimer’s dementia in adults with Down syndrome. Alzheimers Dement (Amst). 2020;12(1):e12044.

45. Igwe KC, Lao PJ, Vorburger RS, et al. Automatic quantification of white matter hyperintensities on T2-weighted fluid attenuated inversion recovery magnetic resonance imaging. Magn Reson Imaging. 2022;85:71–79.

46. Gutierrez J, Elkind MSV, Dong C, et al. Brain Perivascular Spaces as Biomarkers of Vascular Risk: Results from the Northern Manhattan Study. AJNR Am J Neuroradiol. 2017;38(5):862–867.

47. Wardlaw JM, Valdes Hernandez MC, Munoz-Maniega S. What are White Matter Hyperintensities Made of? Relevance to Vascular Cognitive Impairment. J Am Heart Assoc. 2015;4(6).

48. Gouveia-Freitas K, Bastos-Leite AJ. Perivascular spaces and brain waste clearance systems: relevance for neurodegenerative and cerebrovascular pathology. Neuroradiology. 2021;63(10):1581–1597.

49. Helman AM, Siever M, McCarty KL, et al. Microbleeds and Cerebral Amyloid Angiopathy in the Brains of People with Down Syndrome with Alzheimer’s Disease. Journal of Alzheimer’s disease : JAD. 2019;67(1):103–112.

50. Nakata-Kudo Y, Mizuno T, Yamada K, et al. Microbleeds in Alzheimer disease are more related to cerebral amyloid angiopathy than cerebrovascular disease. Dementia and geriatric cognitive disorders. 2006;22(1):8–14.

51. Xu WH. Large artery: an important target for cerebral small vessel diseases. Ann Transl Med. 2014;2(8):78.

52. Breteler MM. Vascular risk factors for Alzheimer’s disease: an epidemiologic perspective. Neurobiol Aging. 2000;21(2):153–160.

53. Kennelly SP, Lawlor BA, Kenny RA. Blood pressure and the risk for dementia: a double edged sword. Ageing Res Rev. 2009;8(2):61–70.

54. Carson AP, Howard G, Burke GL, Shea S, Levitan EB, Muntner P. Ethnic differences in hypertension incidence among middle-aged and older adults: the multi-ethnic study of atherosclerosis. Hypertension. 2011;57(6):1101–1107.

55. Avolio AP, Kuznetsova T, Heyndrickx GR, Kerkhof PLM, Li JK. Arterial Flow, Pulse Pressure and Pulse Wave Velocity in Men and Women at Various Ages. Adv Exp Med Biol. 2018;1065:153–168.

